# Racial and Ethnic Disparities in COVID-19 Infection and Mortality in the United States: A state-wise update

**DOI:** 10.1101/2020.12.03.20243360

**Authors:** Zhaoying Xian, Anshul Saxena, Zulqarnain Javed, John E. Jordan, Safa Alkarawi, Safi U Khan, Karan Shah, Farhaan Vahidy, Khurram Nasir, Prachi Dubey

## Abstract

**Objectives:** To evaluate COVID-19 infection and mortality in ethnic and racial sub-groups across all states in the United States.

**Methods:** Publicly available data from “The COVID Tracking Project at The Atlantic” was accessed between 09/09/2020 and 09/14/2020. For each state and the District of Columbia, % infection, % death, % population proportion for subgroups of race (African American (AA), Asian, American Indian or Alaska Native, (AI/AN) and White), and ethnicity (Hispanic/Latino, and non-Hispanic), were recorded. Absolute and relative excess infection (AEI and REI) and mortality (AEM and REM) were computed as absolute and relative difference between % infection or % mortality and % population proportion for each state. Median (IQR) REI is provided below.

**Results:** The Hispanic population had a median of 158% higher COVID-19 infection relative to their % population proportion (median REI 158%, [IQR: 100% to 200%]). This was followed by AA, with 50% higher COVID-19 infection relative to their % population proportion (median REI, 50% [IQR 25% to 100%]). The AA population had the most disproportionate mortality with a median of 46% higher mortality than % population proportion, (median REM 46% [IQR, 18% to 66%]). Disproportionate impact of COVID-19 was also seen in AI/AN and Asian population with ≥100% excess infections than % population proportion seen in 35 states for Hispanic, 14 states for AA, 9 states for AIAN, and 7 states for Asian populations. There was no disproportionate impact in the white population in any state.

**Conclusions:** Racial/ethnic minorities (AA, Hispanic, AIAN and Asian populations) are disproportionately affected by COVID 19 infection and mortality across the nation. These findings underscore the potential role of social determinants of health in explaining the disparate impact of SARS-CoV-2 on vulnerable demographic groups, as well as the opportunity to improve outcomes in chronically marginalized populations.

## Introduction

Since January 2020, when the first Coronavirus disease 2019 (COVID-19) positive patient was identified in the United States, there have been over 13 million reported cases and over 260 thousand reported fatalities [1]. Although COVID-19 can affect all ages regardless of medical comorbidities, hospitalized patients are more likely to be older and demonstrate at least 1 medical comorbidity [2]. It has also become increasingly evident that in addition to traditional risk factors, there are widespread racial and ethnic disparities in COVID-19 infection and mortality rates, with disproportionately higher impact in vulnerable racial and ethnic minority populations [3–7]. A recent study based on US Department of Veterans Affairs data showed an excess burden of SARS-CoV-2 infection in African American (AA) and Hispanic populations that was not entirely explained by underlying medical conditions, residence, or site of care; however, the study did not find disparities in excess 30-day mortality across racial and ethnic subgroups [8]. This observation however is not aligned with a prior Centers for Disease Control and Prevention (CDC) report, which showed poor outcomes in the AA population with 1.4 times increased risk of hospitalization, twice the risk of ICU care or ventilator support, and 1.36 times increased risk of death compared to Whites [9]. Gross et al, found a robust association between AA race, Latinx ethnicity, and estimated age adjusted COVID-19 mortality [3]. Overall mortality was highest in AAs with 1 in 1020 faced death attributed to COVID-19, followed by Indigenous Americans, Pacific Islanders and Latinos [3]. In addition, a study based on community level disparities in urban US counties in large metropolitan areas showed excess infections and mortality that are not explained by income, bolstering the complex causal pathways that culminate into these striking disparities [10].

Given the demographic diversity of the US population, and the unique challenges faced by underserved racial/ethnic minorities in each state, it is imperative to examine the extent of disparities in COVID-19 prevalence and overall mortality by race and ethnicity on a national scale in a state-wise manner. Further, the racial/ethnic composition of hospitalized patients might differ considerably from that of the general population. In this context, evidence from population-based studies is critical from a quality of care, resource distribution and utilization, and health outcomes standpoint. Consequently, we evaluated the extent of racial and ethnic disparities in COVID-19 infection and mortality as a function of racial/ethnic composition of the general population across all US states and the District of Columbia.

## Materials and methods

### Data source

We used publicly available data from “The COVID Tracking Project at The Atlantic” between September 10, 2020 to September 14, 2020 for this study (https://covidtracking.com/race/dashboard). The COVID Tracking Project at The Atlantic’s data and website content is published under a Creative Commons CC BY 4.0 license, (CC BY 4.0). The COVID Tracking Project derived data regarding racial and ethnic composition of U.S. states and the District of Columbia from United States Census Bureau’s 2018 5-Year estimates. For each state and Washington D.C., the percentage of each racial and ethnic category in the population (i.e. *population proportion*), percent of COVID-19 infections or mortality in each racial and ethnic subgroup (# COVID-19 infections or deaths in each category/Total # COVID-19 infections or deaths) were recorded. The race was categorized into African American, (AA), Asian, American Indian or Alaska Native (AI/AN), and White. Ethnicity was characterized into Hispanic/Latino, and Non-Hispanic. We excluded states that were labeled as “other”, that combined racial or ethnic subgroups, such as combining Asian, Pacific Islander, and Native Hawaiian as a pan-racial group or where the numbers were reported as <1%.

### Patient and public involvement

There was no direct patient or public involvement in the design and conduct of this study.

### Statistical analysis

State-wise absolute excess infection (AEI) and mortality (AEM) was calculated as a difference in percentage points between % infection or mortality in each racial and ethnic subgroup and % population proportion of the same racial/ethnic subgroup, (as noted above, % COVID-19 infection or mortality in each group is the number of infection or deaths in each group/total number of COVID-19 infections or deaths). In order to account for differences in population size for racial/ethnic subgroups, relative excess infection (REI) and mortality (REM) rates were calculated as absolute excess infection (AEI) and mortality (AEM) normalized by population proportion of each racial and ethnic subgroup in the population based on respective census estimates for each state. Absolute and relative excess infection and mortality rates were reported as median and interquartile ranges (IQR). For analytic purposes, percentage estimates less than 1% were excluded from analyses.

Heat maps depicting AEI, AEM, REI and REM by race and ethnicity were generated for all 50 states and Washington DC. Missing data on race or ethnicity for a given state (due to low sample size of less than 1%, indistinct classification or lack of reporting) appeared as a missing value on the heat map.

## Results

Race/ethnicity data were available for between 0-100% of cases and of deaths in each state. 42 states (including Washington D.C.) reported race/ethnicity data for at least 66% of their cases. 46 states (included Washington D.C.) reported race/ethnicity data for at least 75% of their deaths. New York did not report race/ethnicity data for cases. Hawaii and North Dakota did not report race/ethnicity data for deaths either. Wyoming only reported non-mutually-exclusive categories for race/ethnicity and was therefore excluded.

### Racial/Ethnic Disparities in COVID-19 Infection

COVID-19 infection was disproportionately high in the Hispanic, AA, AI/AN and Asian American populations, relative to their population proportion in many states. The Hispanic population experienced the highest AEI and REI, followed by AAs (Figure 1A and 1B). The Hispanic population had a median AEI of 14%, and IQR of 8% to 19%, and the AA population had a median AEI of 5%, and IQR of 3% to 9%. Similarly, both Hispanic and AA populations experienced higher REI. The Hispanic population had a median REI of 158%, and IQR 100% to 200%, in contrast to the AA population median REI of 50%, and IQR 25% to 100 %. The Hispanic population experienced ≥100% REI in 35 states while the AA population experienced ≥100% REI in 14 states across the nation. In contrast, median AEI in the white population was −18% and the IQR −23% to −13%, and median REI was −26%, and the IQR −39% to −17% respectively. Disproportionate impact of COVID-19 was also seen in AI/AN and Asian populations in many states (Figure 1A and 1B), with 100% or more REI in 9 states for AI/AN populations, and in 7 states for Asian populations.

**Figure 1A:**
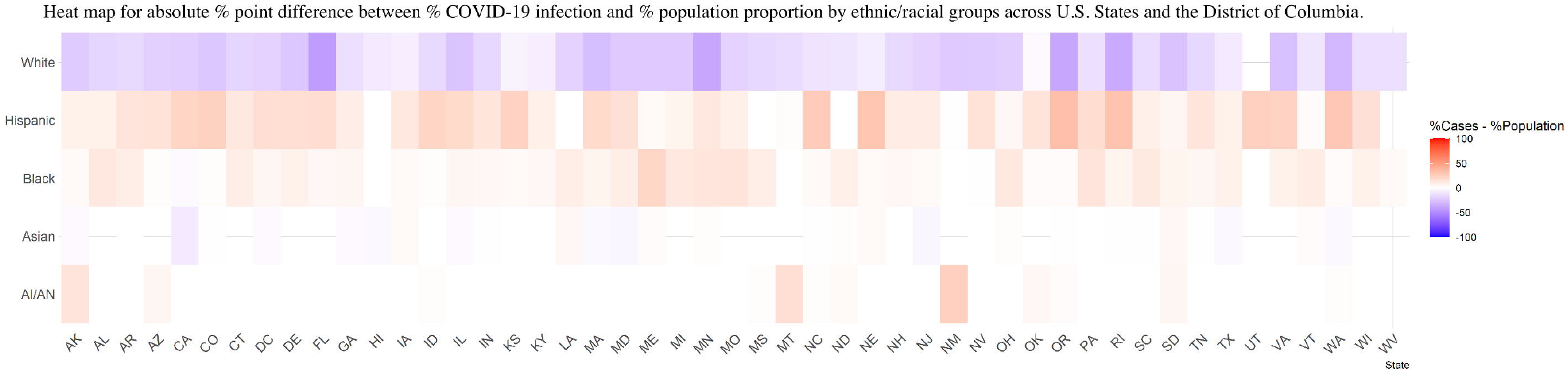
Heat map showing absolute percentage point difference between % COVID-19 infection and % population proportion by ethnic/racial groups across U.S. States and the District of Columbia.

**Figure 1B:**
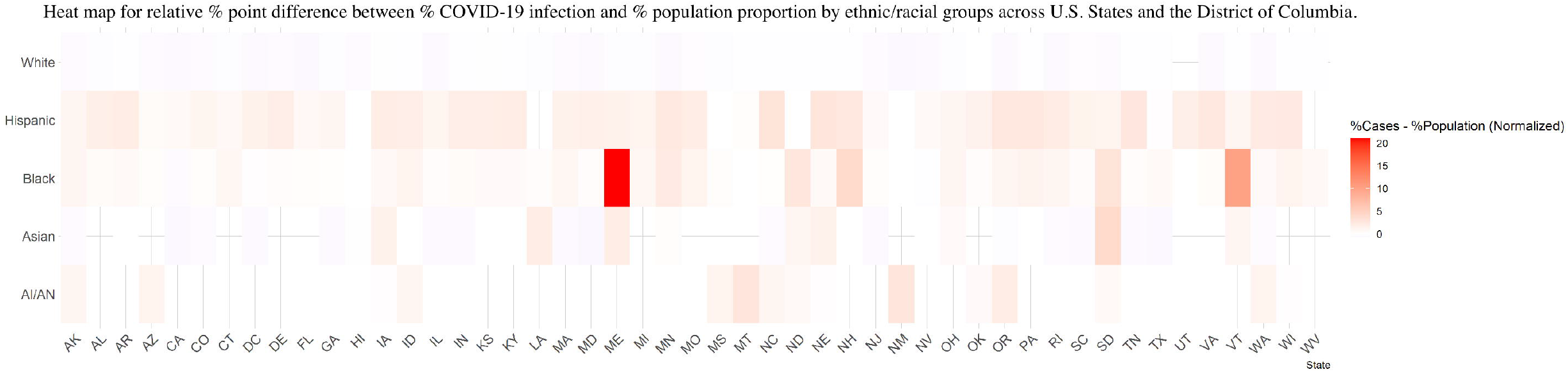
Heat map showing relative percentage point difference between % COVID-19 infection and % population proportion by ethnic/racial groups (normalized by % population proportion) across U.S. States and the District of Columbia.

### Racial/Ethnic Disparities in COVID-19 Mortality

African American populations had disproportionately highest COVID-19 mortality (Figures 1C, Figure 1D). The African American population had an AEM median of 5% (IQR: 1% to 11%), and REM median of 46% (IQR: 18% to 66%). Hispanic, White, AI/AN and Asian populations showed 0 or <0 median AEM or and REM. However, some states showed disparities in mortality even in these subgroups, such as the Hispanic population with a 17% AEM and 43% REM in Texas, while the AI/AN subgroups had a 45% AEM and 500% REM in New Mexico. The Asian American population had a 4% AEM and 50% REM in Nevada.

**Figure 1C:**
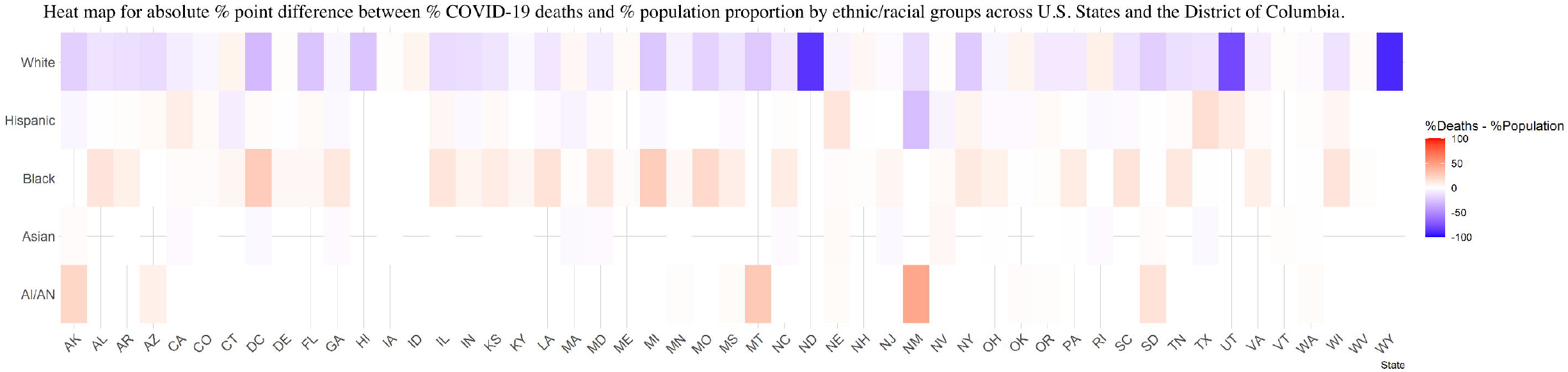
Heat map showing absolute percentage point difference between % COVID-19 deaths and % population proportion by ethnic/racial groups across U.S. States and the District of Columbia.

**Figure 1D:**
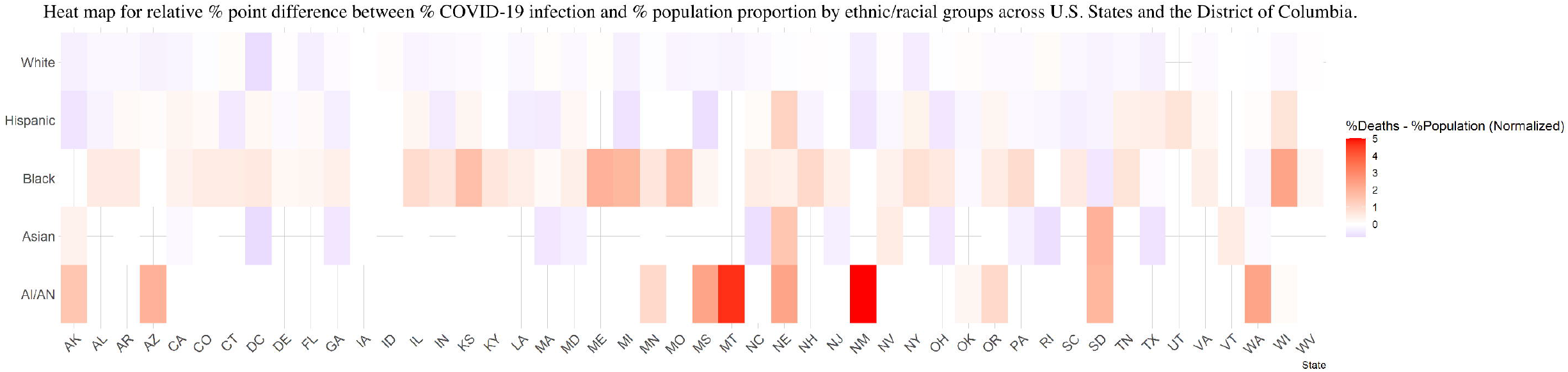
Heat map showing relative percentage point difference between % COVID-19 deaths and % population proportion in ethnic/racial groups (normalized by % population proportion) across U.S. States and the District of Columbia.

## Discussion

We found widespread racial and ethnic disparities in COVID-19 infection and mortality. We summarized these findings in a state-wise manner across the United States. Our findings are consistent with previous similar reports [3–7,11].

Hispanic and AA populations have most disproportionately higher COVID-19 infection and AA population has most disproportionately higher COVID-19 related mortality across the nation. AI/AN and Asian American population also showed disproportionately higher infection and mortality in some states. Although this is susceptible to underestimation given many states with less than 1% representation for these minority subgroups were excluded from analyses. In addition, the excess mortality estimates in our study are not age adjusted, thus they are likely an underestimate relative to other previously published age adjusted estimates, [3].

While we do not have specific information on social determinants of health (SDOH), to understand the exact etiologies of these disparities, we infer a significant impact of SDOH in driving these disparities based on vast body of prior evidence indicating impact of SDOH mediated via environment, occupation, and access to care, and higher burden of co-morbidities in chronically marginalized populations [12–14]. The higher burden of co-morbidities are known to adversely impact disease severity in COVID-19 [15].

Higher levels of obesity, in particular, may result in a chronic, pro-inflammatory state that compounds the cytokine release syndrome observed in COVID-19 [16] [17]. SDOH such as food insecurity – with underlying economic instability (low SES, unstable jobs) result in suboptimal nutrition from inexpensive diets (e.g., high sugar, fats, or high salt) – acting as predictors of diabetes, hypertension and obesity in racial/ethnic minorities. [18], The impact of these factors can be further exacerbated by lack of access to quality and timely care in these minority population subgroups [6,19,20].

Minority populations rely on government funded healthcare facilities, which often have chronically under-resourced infrastructures, not equipped to mobilize at an operational scale needed for a rapidly spreading and potentially lethal infection like COVID-19. This is particularly relevant for the AI/AN population, who rely heavily on the Indian Health Service (IHS) to provide medical care in tribal reservations, including the Navajo Nation, the largest reservation severely impacted by COVID-19. IHS facilities are unable to respond to a crisis of this magnitude due to limited hospital beds and ICU’s. In addition, IHS facility may be inaccessible to those in remote areas and IHS does not cover care provided by non-IHS providers or facilities; a critical factor that was not addressed in the funds provided by the CARES act stimulus bill [5].

Communication challenges due to limited english proficiency and lower health literacy results in additional barriers in seeking and adhering to treatment in minority populations [21–23]. Furthermore, these groups are more likely to be “front-line workers” (e.g., grocery, public transport, or health-care employees) with greater in person interactions and without the flexibility of remote employment [24,25]. It is important to note that socio-economic differences are not a singular explanation for disparities. For example, a recent study found use of public transportation as an independent predictor of COVID-19 disparities even after adjusting for income, poverty rates, education, occupation or access to insurance in AA and AIAN populations, [26]. Commuting to work in public transportation and living in crowding households is not conducive for physical distancing, which is critical for mitigating exposure to a contagious respiratory illness such as COVID-19 [5–7].

Our study has several limitations, mostly stemming from inconsistent and incomplete reporting of race/ethnicity data. There were inconsistencies in race/ethnic categorization across states. The categories were not mutually exclusive, with overlap between race and ethnicity. Many states don’t clarify the composition of the category labeled as “Others”, or may include Asian as “pan-racial,” effectively combining Asian, Pacific Islander and Native American subgroups. Some states reported race/ethnicity data on only a minority of cases at the time of data query. Some groups such as AI/AN and Asian populations had low representation in many states, (<1%) and had to be excluded from analyses. We do not have detailed clinical and socio-demographic information to precisely understand the impact of SDOH, age and other risk factors.

In conclusion, African American, Hispanic, AI/AN, and Asian American populations are disproportionately impacted by COVID-19 both in terms of burden of infection and mortality. We provide a state-wise summary of COVID-19 associated disparities across the United States. Hispanics and AAs tend to experience the greatest disparities in infection while AAs tend to have the greatest disparities in mortality nationwide. We also observed higher infection and mortality in AI/AN and Asians in some states. We acknowledge that the assessment of disparities in groups such as in AI/AN and Asians can be underestimated in states with <1% population representation of these group and that lack of age adjusted mortality estimates can underestimate mortality in groups with younger populations such as the Hispanic population.

Our findings reinforce the urgent need for targeted interventions to address COVID-19 infection and mortality disparities tailored to the needs of states based on the specific pattern of disparity burden. This is key to ensure the minorities in specific states are adequately represented in resource allocation, and implementation of public health and policy measures. It is also imperative to address underlying social determinants of health including access to care, health literacy, housing environments, nutrition, and foster continuous future access to preventive care.

## Data Availability

All of the data reported in the manuscript is available through COVID-19
The COVID Tracking Project at The Atlantic https://covidtracking.com/race/dashboard
The data is periodically updated on the website. We have stored the relevant data accessed in 09/10/2020 and 09/14/2020 for this study.

## Acknowledgements

We appreciate the “The COVID Tracking Project at The Atlantic” for making the race/ethnicity data on COVID-19 publicly available, https://covidtracking.com/race/dashboard. The COVID Tracking Project at The Atlantic’s data and website content is published under a Creative Commons CC BY 4.0 license, (CC BY 4.0).

